# Generative Deep Learning Maps Brain Oxygen Metabolism and Its Age-Related Decline from Two Standard Noninvasive MRI Scans

**DOI:** 10.64898/2026.11.17.26368289

**Authors:** Gabriel Castrillon, Pietro Marcolongo, Urmi Bhattacharyya, Samira M. Epp, Antonia Bose, Stephan Kaczmarz, Gabriel Hoffmann, Elisa Saks, Daniel Rueckert, Moritz Zaiss, Christine Preibisch, Valentin Riedl

## Abstract

**Purpose:** To develop a generative deep learning model mapping the cerebral metabolic rate of oxygen (CMRO_2_) from minimal, contrast-free MRI.

**Materials and Methods:** This retrospective study used 3-T MRI of healthy adults from one center (May 2015-June 2025). A conditional generative adversarial network (GAN) was trained to reproduce contrast-based analytical CMRO_2_ from progressively fewer quantitative MRI inputs (cerebral blood flow [CBF], T2*, T2, cerebral blood volume), T1-weighted MRI, or resting-state functional MRI. Models were trained on 54 participants and tested on 41 (25 women; mean age, 49 years ± 23 [SD]), on calculation-task data, and, without retraining, on same-protocol (n = 24) and cross-vendor (n = 12) data. Agreement (r, mean absolute error [MAE]) was compared with the test-retest error of analytical CMRO_2_, and repeatability (n = 9) with intraclass correlation coefficients (ICCs).

**Results:** A GAN using only CBF and T2* agreed with analytical CMRO_2_ as well as one adding T2 (r = 0.89 [95% CI: 0.88, 0.90] for both), with errors below the between-day test-retest error (median MAE, 38.1 vs 46.5 µmol/100 g/min); CBF alone, T1-weighted MRI, and functional MRI performed worse (P ≤ .004). It was as repeatable as analytical CMRO_2_ (median ICC, 0.87 for both), preserved task-evoked patterns (Dice, 0.78), agreed with same-protocol (r = 0.89) and, less, cross-vendor data (r = 0.81; P ≤ .001), and captured the age-related decline (6.8 vs analytical, 5.0 µmol/100 g/min per decade).

**Conclusion:** Generative deep learning mapped brain oxygen metabolism from two contrast-free scans with repeatability comparable to the reference.

## Introduction

With increasing life expectancy, brain disorders are rising worldwide (1), and multi-omics, neuroimaging, and transcriptomic studies link accelerated brain aging in the early 40s and 60s to energy metabolism (2,3). In Alzheimer disease, which affects one-third of people aged 85 years or older (4), declines in brain energy metabolism may precede cognitive impairment (5), and reduced regional oxygen metabolism is associated with cognition and tau deposition (6,7). Thus, in vivo measures of brain energy metabolism could help distinguish healthy from pathological aging.

The brain depends on a continuous supply of glucose and oxygen. Because the full oxidation of glucose yields about 15 times more energy than glycolysis, the cerebral metabolic rate of oxygen (CMRO_2_) is the most direct marker of cerebral energy production (8). ^15^O-PET, the reference standard for CMRO_2_, requires an on-site cyclotron and arterial blood sampling (9), while contrast-agent-free MRI methods yield only whole-brain values or require gas challenges (10). Multiparametric quantitative blood oxygenation level–dependent (mqBOLD) MRI maps CMRO_2_ voxel-wise via Fick’s principle from cerebral blood flow (CBF), measured with arterial spin labeling (ASL), and oxygen extraction fraction (OEF), derived from T2, T2*, and cerebral blood volume (CBV) (Fig. 1a; 11,12). However, CBV requires a gadolinium-based contrast agent, which, even for the macrocyclic agents used here, could leave trace amounts in the brain (13), favoring contrast-free methods for repeated examinations. Long scan times, complex post-processing, and blood sampling for hematocrit further limit regional CMRO_2_ mapping to specialized centers.

**Figure 1:**
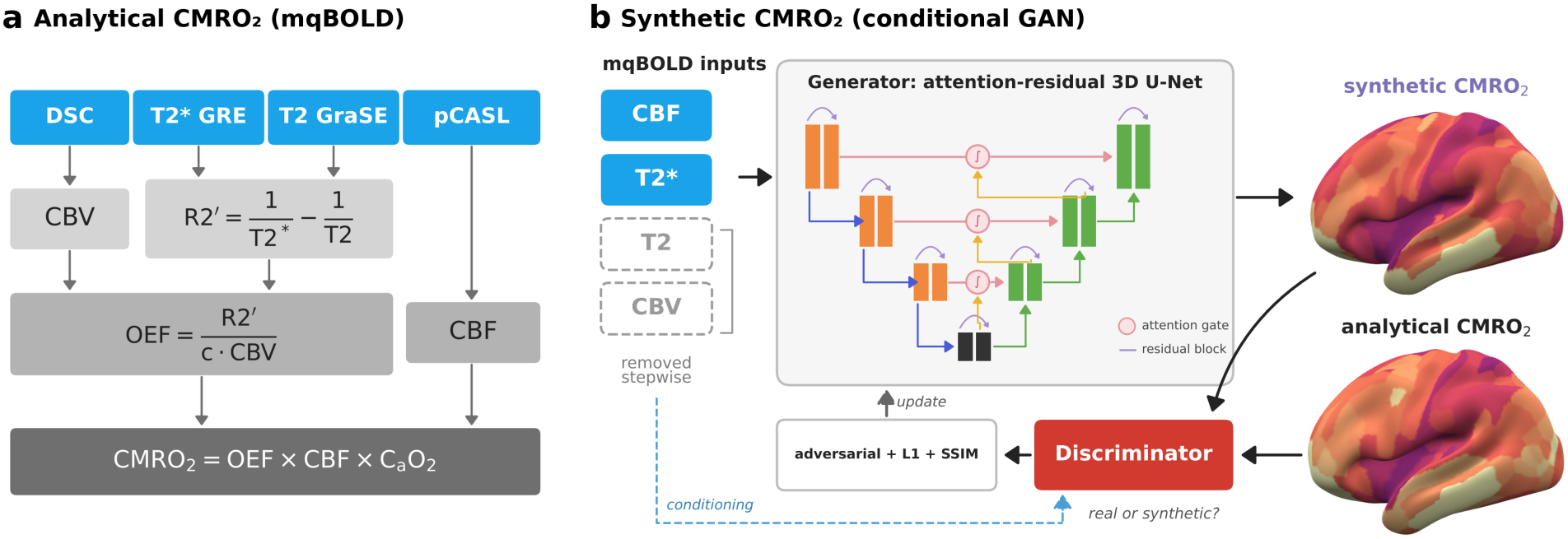
Analytical and synthetic CMRO_2_ mapping. (a) Analytical CMRO_2_ was derived from multiparametric quantitative BOLD MRI using Fick’s principle, as the product of cerebral blood flow (CBF), measured with pseudocontinuous arterial spin labeling, arterial oxygen content (CaO_2_), and oxygen extraction fraction (OEF). OEF was calculated from susceptibility-dependent R2′ values derived from T2 and T2* maps, and cerebral blood volume (CBV) was measured with dynamic susceptibility contrast MRI after contrast agent injection. (b) A conditional generative adversarial network (GAN) synthesized CMRO_2_ from these inputs, which were progressively reduced to CBF and T2*. The generator, a three-dimensional attention-residual U-Net, mapped the inputs to synthetic CMRO_2_, while a discriminator, conditioned on the same inputs, distinguished synthetic from analytical CMRO_2_. Generator weights were updated with the discriminator’s adversarial loss and with L1 and structural similarity losses between synthetic and analytical CMRO_2_. Brain surfaces show group-average analytical and synthetic CMRO_2_. DSC = dynamic susceptibility contrast, GraSE = gradient and spin echo, GRE = gradient echo, pCASL = pseudocontinuous arterial spin labeling, SSIM = structural similarity index measure.

Deep generative models can instead infer missing quantities from available scans. These models translate between MRI contrasts (14), including CBV maps synthesized from ASL (15). They mostly use anatomical T1-weighted images, present in 29 of 35 brain-age studies and 124 of 146 foundation-model datasets, whereas functional MRI (fMRI) is less common (16,17). Resting-state fMRI may nevertheless carry metabolic information, because the degree centrality of brain networks scales with regional energy metabolism (18). However, whether anatomical or functional MRI alone is sufficient to infer quantitative CMRO_2_, and which mqBOLD inputs are required, remains unknown. Moreover, image similarity can arise from anatomy alone, and training-site evaluation inflates performance (19).

Here, we trained a conditional generative adversarial network (GAN) to synthesize CMRO_2_ from progressively reduced MRI inputs, aiming at contrast-free CMRO_2_ mapping in studies of brain aging and metabolic (dys)function (Fig. 1b). We evaluated (i) the minimal input set among mqBOLD inputs (CBV, CBF, T2, and T2*), T1-weighted images, and resting-state fMRI; (ii) repeatability across scans; (iii) sensitivity to task-evoked metabolic changes; (iv) performance in new participants scanned with the same protocol or another vendor; and (v) sensitivity to the age-related decline of CMRO_2_.

## Materials and Methods

### Study Design and Participants

This retrospective model-development study used independently acquired datasets of healthy adults from one center (May 2015-June 2025; Appendix S1). Except for Older-adults II, all were previously reported in studies that did not synthesize CMRO_2_ (Table 1). The local ethics committee approved all studies, and participants gave written informed consent. The data used from these studies were pseudonymized, and high-resolution T1-weighted images were defaced. All healthy participants of the source studies with complete data of sufficient quality were included (for the ongoing Older-adults II study, those scanned by June 30, 2025), except three cross-vendor participants with implausibly high CMRO_2_ (>250 µmol/100 g/min). Sample size was determined by the availability of data. Extended methods are provided in Appendix S1.

**Table 1:**
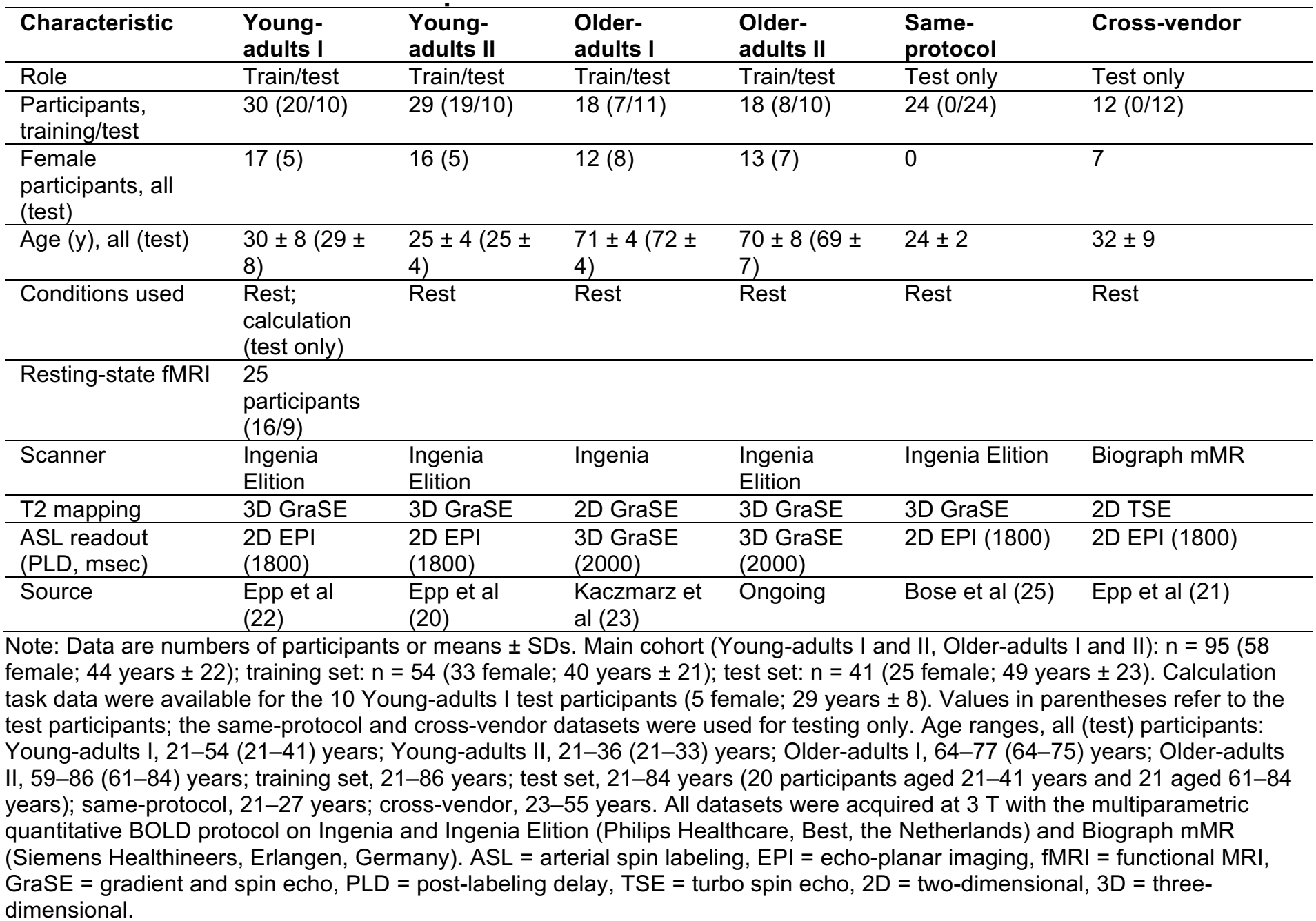
Characteristics and Acquisition Parameters of the Datasets.

| **Characteristic** | **Young-adults I** | **Young-adults II** | **Older-adults I** | **Older-adults II** | **Same-protocol** | **Cross-vendor** |
|---|---|---|---|---|---|---|
| Role | Train/test | Train/test | Train/test | Train/test | Test only | Test only |
| Participants, training/test | 30 (20/10) | 29 (19/10) | 18 (7/11) | 18 (8/10) | 24 (0/24) | 12 (0/12) |
| Female participants, all (test) | 17 (5) | 16 (5) | 12 (8) | 13 (7) | 0 | 7 |
| Age (y), all (test) | 30 ± 8 (29 ± 8) | 25 ± 4 (25 ± 4) | 71 ± 4 (72 ± 4) | 70 ± 8 (69 ± 7) | 24 ± 2 | 32 ± 9 |
| Conditions used | Rest; calculation (test only) | Rest | Rest | Rest | Rest | Rest |
| Resting-state fMRI | 25 participants (16/9) |  |  |  |  |  |
| Scanner | Ingenia Elition | Ingenia Elition | Ingenia | Ingenia Elition | Ingenia Elition | Biograph mMR |
| T2 mapping | 3D GraSE | 3D GraSE | 2D GraSE | 3D GraSE | 3D GraSE | 2D TSE |
| ASL readout (PLD, msec) | 2D EPI (1800) | 2D EPI (1800) | 3D GraSE (2000) | 3D GraSE (2000) | 2D EPI (1800) | 2D EPI (1800) |
| Source | Epp et al (22) | Epp et al (20) | Kaczmarz et al (23) | Ongoing | Bose et al (25) | Epp et al (21) |

Note: Data are numbers of participants or means ± SDs. Main cohort (Young-adults I and II, Older-adults I and II): n = 95 (58 female; 44 years ± 22); training set: n = 54 (33 female; 40 years ± 21); test set: n = 41 (25 female; 49 years ± 23). Calculation task data were available for the 10 Young-adults I test participants (5 female; 29 years ± 8). Values in parentheses refer to the test participants; the same-protocol and cross-vendor datasets were used for testing only. Age ranges, all (test) participants: Young-adults I, 21–54 (21–41) years; Young-adults II, 21–36 (21–33) years; Older-adults I, 64–77 (64–75) years; Older-adults II, 59–86 (61–84) years; training set, 21–86 years; test set, 21–84 years (20 participants aged 21–41 years and 21 aged 61–84 years); same-protocol, 21–27 years; cross-vendor, 23–55 years. All datasets were acquired at 3 T with the multiparametric quantitative BOLD protocol on Ingenia and Ingenia Elition (Philips Healthcare, Best, the Netherlands) and Biograph mMR (Siemens Healthineers, Erlangen, Germany). ASL = arterial spin labeling, EPI = echo-planar imaging, fMRI = functional MRI, GraSE = gradient and spin echo, PLD = post-labeling delay, TSE = turbo spin echo, 2D = two-dimensional, 3D = three-dimensional.

Participants in each main-cohort dataset (Table 1) were manually assigned to training and test sets, balanced for age and sex, without inspecting imaging data. The fMRI models used the 25 Young-adults I participants with resting-state fMRI (16 training, 9 test). Repeatability was assessed in 9 Young-adults II test participants using a second baseline rest acquisition in the same session (20).

### Image Acquisition and Preprocessing

All datasets were acquired at 3 T with the mqBOLD protocol (Table 1; 21). Quantitative maps were coregistered to the T1-weighted image (FSL version 6.0), normalized to MNI152NLin2009cAsym space at 4-mm resolution (fMRIPrep version 20.2.3; ANTs version 2.3.3), and kept in physical units, so that models learned absolute CMRO_2_; T1-weighted and fMRI degree centrality maps were z-scored within gray matter (Table S1; 18). Analyses used the conservative cortical gray matter mask of Epp et al (22), because mqBOLD-derived OEF and CMRO_2_ are biased in white matter and in areas with strong magnetic susceptibility effects (12).

### Analytical CMRO_2_ Maps

Analytical CMRO_2_, the reference standard for all models, was calculated in native space using Fick’s principle as the product of OEF, CBF, and arterial oxygen content, derived from individual hematocrit and pulse oximetry (Fig. 1a; 21). OEF was estimated as R2′/(c · CBV), with R2′ = 1/T2* − 1/T2 and leakage-corrected CBV normalized to 2.5% in white matter; CBF was increased by 25% to compensate for background suppression on Philips data (23). Older-adults I used sex-specific reference hematocrit values (women, 0.40; men, 0.45).

### Synthetic CMRO_2_ Model

CMRO_2_ was synthesized with a conditional GAN (Fig. 1b; Fig. S1; Table S1; 14) with a three-dimensional U-Net generator with residual blocks and attention-gated skip connections, trained with L1, adversarial, and structural similarity losses. All models were trained for 750 epochs on data acquired during a baseline rest condition, with augmentation and fourfold participant-level cross-validation, retaining, per fold, the checkpoint and, per model, the fold with the lowest validation generator loss. Models used all four mqBOLD maps (full model), all seven combinations of CBF, T2, and T2* (three single inputs, three pairs, and one triplet), or T1-weighted images or degree centrality; for each number of mqBOLD inputs, the combination with the lowest cross-validated MAE was selected (Table S3).

### Model Evaluation

Throughout, model performance refers to the voxel-wise Pearson correlation coefficient (r) and slope of the linear regression of synthetic on analytical CMRO_2_ and to the mean absolute error (MAE), both on the group median maps (voxel-wise median across participants) and individually, excluding non-positive voxels and six Glasser parcels per hemisphere with outlying analytical CMRO_2_ next to the petrous bone and frontal sinus (194 of 8155 voxels; Appendix S1). The within-session and between-day intraclass correlation coefficients (ICC) for analytical CMRO_2_ (0.89 and 0.61; 20) were converted into expected test-retest errors, 2/√π · SD · √(1 − ICC), with SD as the between-participant SD per voxel for each dataset (Table S2). Test-retest reliability was assessed as the ICC(2,1) of participant-wise medians across 305 Glasser parcels (24) with at least 5 mask voxels. Task-evoked patterns of analytical and synthetic CMRO_2_ were identified with mean-centered partial least squares analysis (3000 permutations and bootstrap resamples), and their overlap (absolute bootstrap ratio > 2) was quantified with the Dice coefficient.

Age effects were assessed in the test set as the correlation and change per decade of median gray matter CMRO_2_ (Table S4). For age calibration, each training participant, a validation participant in exactly one fold, was predicted by that fold’s model; in these 54 predictions, the error (synthetic minus analytical CMRO_2_) was fitted as a + b · age, and calibrated CMRO_2_ was synthetic CMRO_2−_ (a + b · age). Because the participant-wise regression slope of synthetic on analytical CMRO_2_ also changed with age, age effects were additionally estimated with the slope as a covariate, which requires analytical CMRO_2_. Age slopes were compared by regressing the participant-wise difference on age, equivalent to the age-by-method interaction in a mixed model with a random intercept per participant.

### Statistical Analysis

The 95% CIs were estimated with bias-corrected and accelerated bootstrapping for r and with participant bootstrapping for age slopes (2000 resamples each). Metrics and parcel-wise ICCs were compared between models and conditions using Wilcoxon signed-rank tests with false discovery rate correction, and between the test set and the same-protocol and cross-vendor datasets using Games-Howell tests; Δ denotes the mean paired difference (Wilcoxon tests) or the difference in means (Games-Howell tests). P < .05 indicated statistical significance. Analyses used Python (version 3.13.5), SciPy (version 1.16.1), scikit-learn (version 1.7.1), pyls (commit d8a19d5), and Nilearn (version 0.11.1); deep learning libraries are listed in Table S1.

## Results

### Characteristics of the data cohorts

The main cohort comprised 95 healthy adults with complete data of sufficient quality (58 women; 44 ± 22 years; training, n = 54; test, n = 41); calculation task data were available for 10 test participants, and the same-protocol and cross-vendor datasets comprised 24 and 12 participants, respectively (Table 1).

### CBF and T2* form the minimal input set for CMRO_2_ synthesis

To identify the minimal input set, we compared analytical CMRO_2_, calculated from all four mqBOLD inputs, with synthetic CMRO_2_ from the full model and, for each smaller number of inputs, from the input combination with the lowest cross-validated MAE (Table S3; Fig. 2). At the group level, the full model agreed excellently with analytical CMRO_2_ (r = 0.92 [95% CI: 0.91, 0.93]; slope = 1.08; MAE = 12.7 µmol/100 g/min; Fig. 2b), and agreement remained high without CBV (r = 0.89 [95% CI: 0.88, 0.90]; slope = 0.91; MAE = 16.0; Fig. 2c) and without CBV and T2 (i.e., CBF&T2*: r = 0.89 [95% CI: 0.88, 0.90]; slope = 0.88; MAE = 17.9; Fig. 2d) but decreased with CBF alone (r = 0.79; slope = 0.59; MAE = 22.0; Fig. 2e). Individually, the full model performed best; the without-CBV and CBF&T2* models performed equally in r and MAE (P ≥ .12), with a slightly higher slope for the CBF&T2* model (Δslope = 0.02; P = .005), and both outperformed the CBF model (P < .001; Fig. 2h-2j). Median individual MAEs approached the within-session test-retest error of analytical CMRO_2_ for the full model (25.7 vs 24.7 µmol/100 g/min) and remained below its between-day error for the CBF&T2* model (38.1 vs 46.5; Table S2). Thus, T2 added little information beyond CBF and T2*, which together formed the minimal input set.

**Figure 2:**
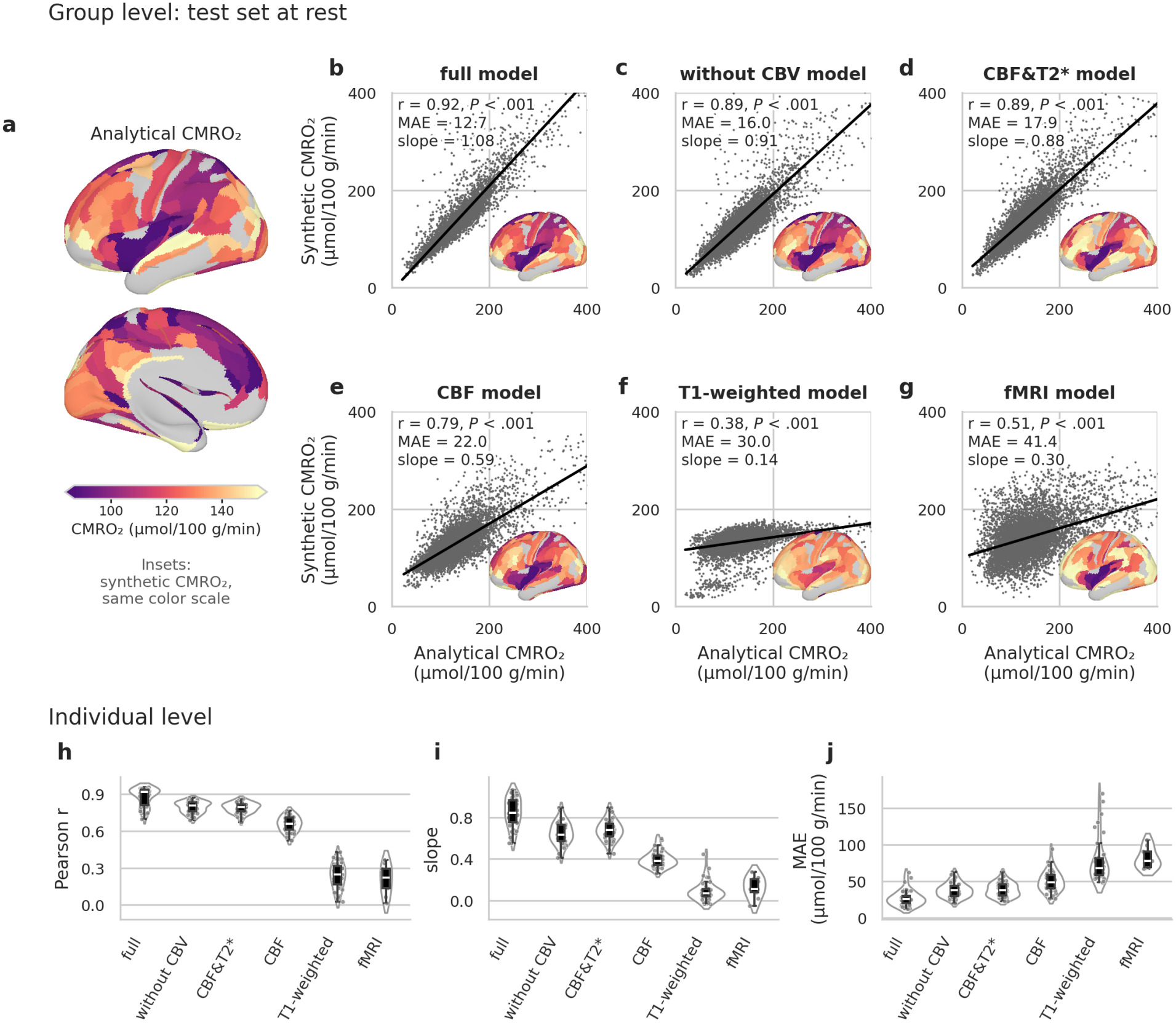
Synthetic CMRO_2_ from reduced inputs at rest. Group level, test set (n = 41): (a) analytical CMRO_2_ as the voxel-wise median across participants, summarized as the median within each Glasser parcel and shown on the inflated left hemisphere (lateral and medial views); (b-g) analytical versus synthetic CMRO_2_ in each voxel of the group median maps (median across participants in each voxel; 7961 gray matter voxels; non-positive values excluded per participant) for the full model (CBF, CBV, T2, and T2*; b), the best-performing models after removing CBV (without CBV; c), CBV and T2 (CBF&T2*; d), and CBV, T2, and T2* (CBF; e), and the single-input models based on T1-weighted MRI (f) and resting-state functional MRI degree centrality (fMRI; n = 9; g). Lines indicate least-squares fits. Insets show the synthetic group median map of each model; the color scale spans the 10th to 90th percentile of the analytical parcel values and is shared by the analytical and synthetic cortical surface maps; parcels excluded from the evaluation or with less than 25% of their voxels or fewer than 5 voxels inside the gray matter mask are shown in gray. Individual level: participant-wise (h) Pearson correlation coefficient (r) and (i) slope of the linear regression of synthetic vs analytical CMRO_2_, and (j) mean absolute error (MAE) for each model in b–g; each dot represents one test participant, boxes show the interquartile range, and white lines the median. CMRO_2_ and MAE values are in µmol/100 g/min.

To test whether anatomical or functional MRI could replace mqBOLD inputs, we trained models on T1-weighted images and resting-state fMRI degree centrality. The T1-weighted model agreed weakly with analytical CMRO_2_ (r = 0.38; slope = 0.14; MAE = 30.0; Fig. 2f), and the fMRI model (n = 9) captured the regional pattern better (r = 0.51; slope = 0.30; MAE = 41.4; Fig. 2g); individually, both performed worse than the CBF model in all metrics (ΔMAE = 27.3 and 32.8 µmol/100 g/min; P < .001 and P = .004; Fig. 2h-2j). Thus, neither anatomical nor functional MRI could replace mqBOLD inputs, and subsequent analyses used the CBF&T2* model.

### Synthetic CMRO_2_ is repeatable and preserves task-evoked metabolic changes

To assess repeatability, we compared the parcel-wise test-retest reliability of synthetic and analytical CMRO_2_ in 9 participants scanned twice within one session (Fig. S2). The full, without-CBV, and CBF&T2* models were as reliable as analytical CMRO_2_ (P ≥ .53; CBF&T2*: median ICC, 0.87 [interquartile range, 0.77–0.92] vs 0.87 [0.78–0.93]), whereas the CBF model was less reliable (0.84 [0.72–0.90]; P = .004).

To test whether models trained on baseline rest data capture task-evoked metabolic changes, we applied them to calculation task data (Fig. 3). During calculation, individual agreement was consistent with rest, but MAEs were slightly higher (ΔMAE = 1.3–2.0 µmol/100 g/min; P ≤ .04) and the correlation of the full model was slightly lower (Δr = −0.005; P = .008; Fig. 3a-3c). Task-evoked patterns, identified separately in analytical and synthetic CMRO_2_ with partial least squares analysis, overlapped closely for the full, without-CBV, and CBF&T2* models (Dice = 0.78–0.81) but less for the CBF model (Dice = 0.59; Fig. 3d-3g). The CBF&T2* model also retained group-level agreement during calculation (r = 0.89; slope = 0.84; MAE = 25.5; Fig.3h). Thus, synthetic CMRO_2_ retained task-evoked metabolic changes, although trained on rest data only.

**Figure 3:**
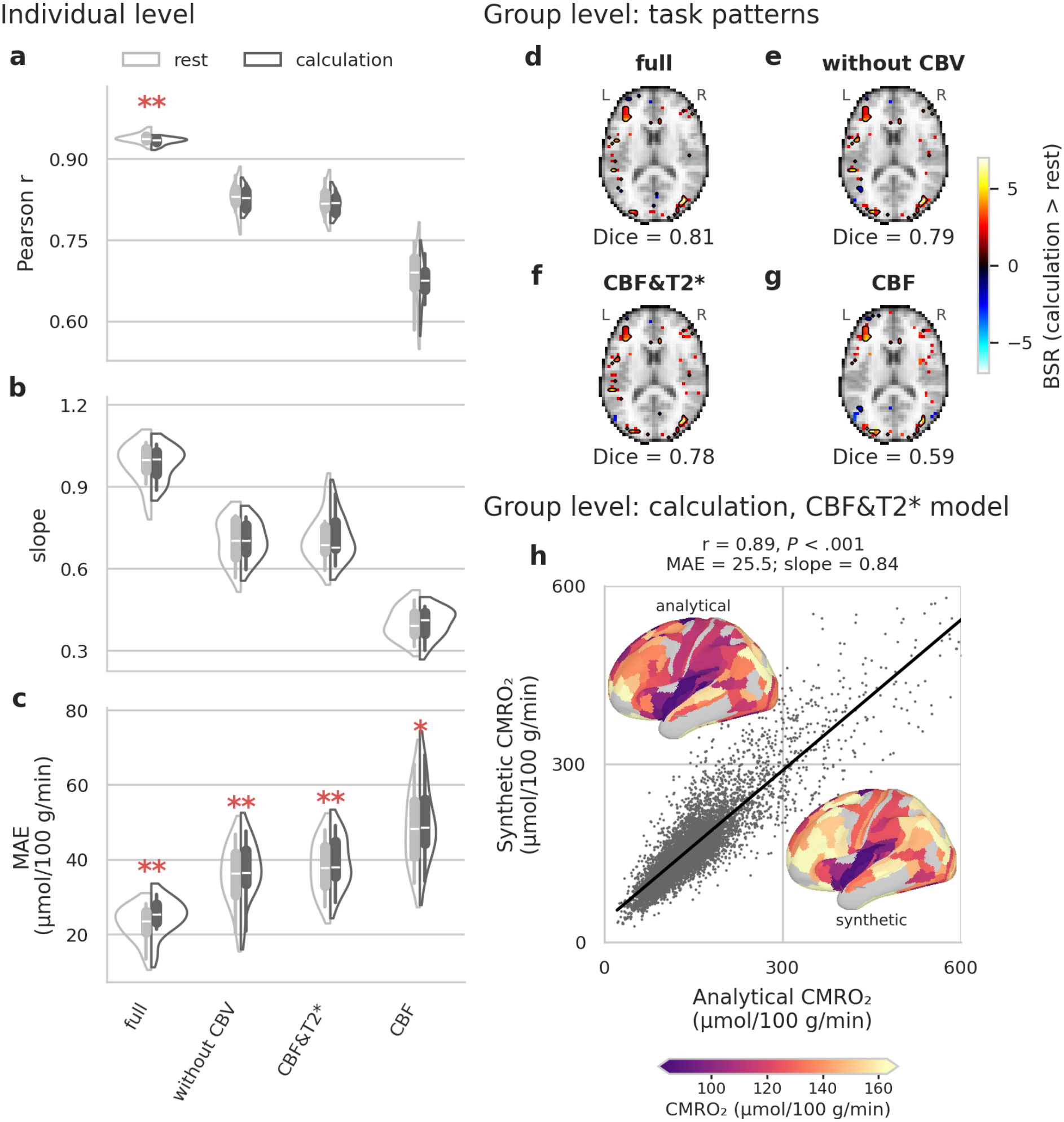
Synthetic CMRO_2_ preserves task-evoked metabolic changes. Rest-trained models were applied to calculation-task data from the Young-adults I test participants (n = 10). Individual level: participant-wise (a) Pearson correlation coefficient (r) and (b) slope of the linear regression of synthetic vs analytical CMRO_2_, and (c) mean absolute error (MAE) at rest (light gray) and during calculation (dark gray). * P < .05, ** P < .01, calculation versus rest (Wilcoxon signed-rank test, false discovery rate corrected across models within each metric). (d-g) Group level: axial slice (z = 15 mm) of the bootstrap ratios (BSRs) of the first latent variable of mean-centered partial least squares analysis contrasting calculation with rest, computed on the synthetic CMRO_2_ of the full (d), without-CBV (e), CBF&T2* (f), and CBF (g) models (|BSR| > 2; positive, calculation > rest; negative, rest > calculation). Black contours outline voxels with |BSR| > 2 in both the synthetic and the analytical CMRO_2_ analyses. Dice coefficients quantify the overlap of voxels with |BSR| > 2 between the synthetic and analytical analyses, irrespective of sign. Background, MNI152NLin2009cAsym template (4 mm). (h) Group level during calculation, minimal-input CBF&T2* model: voxel-wise analytical versus synthetic CMRO_2_ of the group median maps; the line indicates the least-squares fit. Insets show analytical (upper left) and synthetic (lower right) CMRO_2_ as parcel medians on the left lateral surface; the color scale (horizontal bar) spans the 10th to 90th percentile of the analytical parcel values and is shared by the analytical and synthetic maps; parcels with less than 25% of their voxels or fewer than 5 voxels inside the gray matter mask are shown in gray. CMRO_2_ and MAE values are in µmol/100 g/min.

### Synthetic CMRO_2_ transfers to new participants and another vendor

We applied the CBF&T2* model without retraining to the same-protocol and cross-vendor datasets (Fig. 4a-4h). At the group level, the correlation in the same-protocol dataset matched that in the test set, with a higher slope and MAE (r = 0.89; slope = 0.99; MAE = 26.8 µmol/100 g/min; Fig. 4a) and remained high, though lower, in the cross-vendor dataset (r = 0.81; slope = 0.72; MAE = 40.0; Fig. 4e). Individually, the same-protocol dataset showed a correlation similar to that in the test set (P = .99) and slightly higher slope and MAE (Δslope = 0.05, P = .04; ΔMAE = 6.6 µmol/100 g/min, P = .02; Fig. 4b-4d), possibly because its participants were young (21–27 years), whereas all cross-vendor metrics were worse (Δr = −0.07, P < .001; Δslope = −0.11, P = .001; ΔMAE = 25.1 µmol/100 g/min, P < .001; Fig. 4f-4h). Median individual MAEs approached the between-day test-retest error in the same-protocol dataset (43.0 vs 41.9 µmol/100 g/min) and exceeded it in the cross-vendor dataset (58.9 vs 52.8; Table S2). Thus, the model held in new participants scanned with the training protocol and, with lower agreement, transferred to another vendor with a similar protocol.

**Figure 4:**
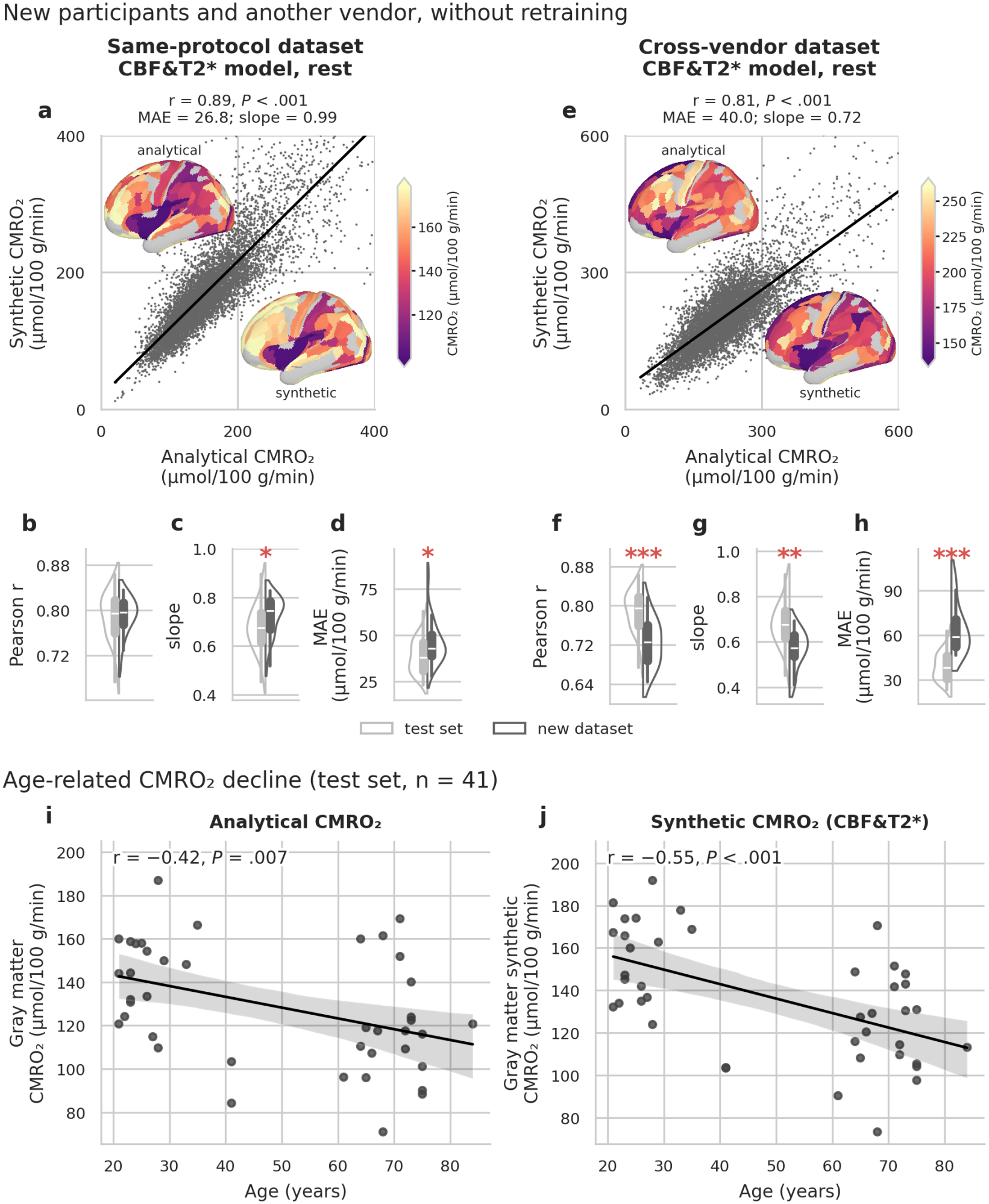
Performance in new participants and across vendors, and age sensitivity of the minimal-input model. The CBF&T2* model was applied without retraining to an independent same-protocol dataset (a-d; n = 24; 21–27 years) and a cross-vendor dataset acquired with another vendor and a similar protocol (e-h; n = 12; 23–55 years). (a, e) Group level: voxel-wise analytical versus synthetic CMRO_2_ of the group median maps; lines indicate least-squares fits. Insets show analytical (upper left) and synthetic (lower right) CMRO_2_ as parcel medians on the left lateral surface; the color scale (vertical bar) spans the 10th to 90th percentile of the analytical parcel values and is shared by the analytical and synthetic maps; parcels with less than 25% of their voxels or fewer than 5 voxels inside the gray matter mask are shown in gray. (b-d, f-h) Individual level: participant-wise Pearson correlation coefficient (r; b, f) and slope (c, g) of the linear regression of synthetic vs analytical CMRO_2_, and mean absolute error (MAE; d, h) in the test set (light gray; n = 41) and in the new dataset (dark gray). * P < .05, ** P < .01, *** P < .001 (Games-Howell test). (i, j) Across the test participants (n = 41), median gray matter analytical CMRO_2_ (i) and uncorrected synthetic CMRO_2_ of the CBF&T2* model (j) decreased with age; age-calibrated and slope-adjusted declines are reported in the text. Lines indicate least-squares fits with bootstrapped 95% CIs (shaded). CMRO_2_ and MAE values are in µmol/100 g/min.

### Synthetic CMRO_2_ captures the age-related metabolic decline

We related each test participant’s median gray matter CMRO_2_ to age. Analytical CMRO_2_ declined by 5.0 µmol/100 g/min per decade (95% CI: 2.0, 8.0; r = −0.42; P = .007; Fig. 4i), whereas synthetic CMRO_2_ declined by 6.8 (95% CI: 4.0, 9.8; r = −0.55; P < .001; Fig. 4j). The synthetic decline was steeper by 1.8 µmol/100 g/min per decade (95% CI: 0.4, 3.2; P = .02), reflecting an age-dependent prediction error: synthetic CMRO_2_ overestimated analytical CMRO_2_ more in younger participants, and the participant-wise regression slope also decreased with age (difference: r = −0.37; P = .02; slope: r = −0.31; P = .047; Table S4). For new participants without analytical CMRO_2_, we subtracted the age-specific error estimated by cross-validation in training participants predicted by fold models that had not seen them (age calibration). This reduced the synthetic decline to 6.0 µmol/100 g/min per decade (95% CI: 3.2, 9.1), with no statistically significant difference from the decline observed for analytical CMRO2 (difference, 1.1 µmol/100 g/min per decade [95% CI: −0.4, 2.4]; P = .16). When analytical CMRO_2_ is available, as here, adjusting both age associations for the participant-wise regression slope likewise left no significant difference (1.0 µmol/100 g/min per decade [95% CI: −0.2, 2.1]; P = .13). Thus, synthetic CMRO_2_ captured the age-related decline but exaggerated it before correction.

## Discussion

Regional CMRO_2_ can be synthesized from two noninvasive MRI scans, CBF from ASL and T2* from multi-echo gradient-echo imaging, as accurately as from three mqBOLD inputs, with errors below the between-day test-retest error of the analytical reference method. Critically, synthetic CMRO_2_ preserved physiological variation beyond image similarity, recovering task-evoked metabolic patterns and age-related decline. Compared with mqBOLD, the model maps CMRO_2_ without need for contrast agent, blood sampling, or complex post-processing, using two of the four acquisitions.

In mqBOLD, R2′ is derived from separate T2* and T2 maps and, combined with deoxygenated blood volume, yields OEF. The original quantitative BOLD approach instead fits R2′ and deoxygenated blood volume from dedicated spin-echo-based sequences (26), whereas combined quantitative susceptibility mapping and quantitative BOLD approaches derive OEF from multi-echo gradient-echo data, by model fitting (27) or a neural network (28). Our network inferred CMRO_2_ from CBF and T2* alone, plausibly because T2* depends on both T2 and R2′ and CBV scales with CBF (15,29). In contrast, T1-weighted images carried little CMRO_2_ information, so anatomy alone cannot explain the agreement of the mqBOLD-based models.

Resting-state fMRI captured regional CMRO_2_ better, in line with the scaling of degree centrality with energy metabolism (18), but, as a relative measure without physiological units, degree centrality captured the regional pattern rather than individual absolute values.

Because ASL is recommended for clinical use (30) and T2* can be mapped with widely available multi-echo gradient-echo sequences, the minimal-input model narrows the gap to clinical CMRO_2_ mapping, although clinical protocols, such as those optimized for quantitative susceptibility mapping, will require retraining or fine-tuning. A high ICC reflects repeatability, not accuracy: a model that does not reproduce scan-to-scan noise can be more reliable than its reference without being more accurate. Without retraining, agreement remained high with another vendor and a similar protocol (r = 0.81), a validation on independently acquired data that is often missing for medical imaging models (19). The lower slope (0.72) may partly reflect the reference: T2 is not a model input, but the cross-vendor T2 mapping inflates T2 and thereby analytical CMRO_2_ (12,21), which therefore differed from the training data.

Synthetic CMRO_2_ captured the age-related decline reported with PET (31) but overestimated the analytical decline by about one-third. This reflected age-dependent prediction error, a bias also recognized in brain-age models (32). Both corrections, by the regression slope when analytical CMRO_2_ is available and by age-based calibration otherwise, left no significant difference between the synthetic and analytical declines, but this does not establish equivalence: training errors showed no significant age trend (−0.8 µmol/100 g/min per decade; P = .60), so calibration had a modest effect, and the confidence interval allowed discrepancies of up to 2.4 µmol/100 g/min per decade. Agreement across the lifespan therefore remains to be established in larger cohorts.

Our study has limitations. First, participants were healthy adults from a single center; as CBV and OEF can change independently of CBF in disease (33), patient studies are required. Second, synthetic CMRO_2_ inherits the model assumptions and systematic errors of mqBOLD, which affect absolute accuracy and limit the analysis to cortical gray matter, excluding areas with strong magnetic susceptibility effects (11,12), although mqBOLD OEF agreed with ^15^O-PET (34). Third, the inputs lack individual arterial oxygen content, so hematocrit differences between women and men are not captured. Fourth, at 4-mm resolution, atrophy-related partial volume effects may explain lower spatial agreement in older participants and contribute to the steeper synthetic age decline. Fifth, age coincided with the ASL protocol in the test set (younger participants, two-dimensional readout with a 1800-msec post-labeling delay; older participants, three-dimensional readout with 2000 ms), so the age effects cannot be fully separated from protocol effects. Finally, reliability was assessed within one session; longitudinal studies should assess between-day reliability and test directly acquired R2′ maps (35).

In summary, generative deep learning mapped regional CMRO_2_ and its age-related decline from two standard contrast-free MRI scans, with errors below the between-day test-retest error and repeatability equal to that of the analytical reference. Once validated in patients and across scanners, synthetic CMRO_2_ could bring oxygen metabolism imaging to lifespan cohorts and older adults at risk of dementia.

## Data Availability

Data generated or analyzed during the study are available as follows. The Young-adults I (https://openneuro.org/datasets/ds004873/) and cross-vendor (https://openneuro.org/datasets/ds008786) datasets are publicly available on OpenNeuro. The Young-adults II dataset will be made publicly available upon release of the corresponding preprint (early 2027). The Older-adults I and II datasets and the same-protocol dataset are available from the principal investigators of the source studies upon reasonable request.

https://github.com/NeuroenergeticsLab/synthCMRO2

## Data Sharing

Data generated or analyzed during the study are available as follows. The Young-adults I (https://openneuro.org/datasets/ds004873/) and cross-vendor (https://openneuro.org/datasets/ds008786) datasets are publicly available on OpenNeuro. The Young-adults II dataset will be made publicly available upon release of the corresponding preprint (early 2027). The Older-adults I and II datasets are available upon reasonable request to C.P., and the same-protocol dataset upon request to V.R. The code repository (https://github.com/NeuroenergeticsLab/synthCMRO2) contains the group-average values, the individual statistics, and all information needed to create the figures and to run the complete pipeline on the publicly available data, including tests and the software environment.

## Acknowledgments

We thank Andrzej Liebert for his advice on the design of the generative network architecture. V.R. has been funded by the European Union (ERC Starting Grant, SUGARCODING, 759659) and C.P. by the German Research Foundation (DFG, 563563598). The funders had no role in study design, data collection, analysis, interpretation, or manuscript preparation.

