## Supplementary Material for "Generative Deep Learning Maps Brain Oxygen Metabolism and Its Age-Related Decline from Two Standard Noninvasive MRI Scans"

#### Appendix S1: Extended Methods

##### Participants

We included data from six datasets of healthy participants acquired at the Klinikum rechts der Isar, Technical University of Munich (Table 1). The main cohort comprised four datasets acquired at baseline rest (n = 95; 58 female; mean age, 44 years ± 22 [SD]): Young-adults I (20 training and 10 test participants; 1), Young-adults II (19 training and 10 test participants; 2), Older-adults I (7 training and 11 test participants), and Older-adults II (8 training and 10 test participants). Participants were assigned to either the training set (n = 54; 33 female; 40 years ± 21) or the test set (n = 41; 25 female; 49 years ± 23), so that no participant contributed data to both sets; assignment was done by hand, balancing age and sex between the sets without inspecting imaging data. Data were acquired between May 2015 and May 2017 (Older-adults I), May and November 2020 (Young-adults I), May 2022 and June 2023 (Young-adults II), July 2021 and June 2023 (same-protocol dataset), March 2019 and April 2021 (cross-vendor dataset), and from October 2021 onward (Older-adults II, ongoing; participants scanned by June 30, 2025, were included). The models were further evaluated on three datasets: (i) the Young-adults I test participants during a calculation task (n = 10; 5 female; 29 years ± 8); (ii) an independent dataset acquired with the same vendor and protocol as the training data (same-protocol dataset; n = 24; all male; 24 years ± 2; 3); and (iii) a dataset acquired with another vendor and a similar protocol (cross-vendor dataset; n = 12; 7 female; 32 years ± 9; 4). Resting-state fMRI data were available for 25 Young-adults I participants (16 training, 9 test). Repeatability was assessed in 9 Young-adults II test participants with a second baseline rest acquisition in the same session (2). Only healthy participants with complete data of sufficient quality, as assessed in the source studies, were included; participants with contrast-agent issues, poor data quality, or brain lesions were excluded (1), as were three cross-vendor participants with implausibly high $CMRO_2$ (>250 µmol/100 g/min). All participants were free of neurological and psychiatric conditions and gave written informed consent, and the local ethics committee approved all studies, which were conducted in accordance with the Declaration of Helsinki. The data used were pseudonymized, and high-resolution T1-weighted images were defaced.

##### Image Acquisition

The main cohort and the same-protocol dataset were acquired on 3-T Ingenia or Ingenia Elition scanners (Philips Healthcare, Best, the Netherlands) with a 32-channel head coil; dataset-specific parameters are listed in Table 1. The multiparametric quantitative BOLD (mqBOLD) protocol (5–7) comprised four acquisitions. T2 mapping used a multi-echo three-dimensional gradient and spin echo (GraSE) sequence (two-dimensional in Older-adults I; 8 echoes; first echo time and echo spacing, 16 msec; repetition time, 251 msec; flip angle, 90°; voxel size, 2 × 2 × 3.3 mm³; 35 sections). T2* mapping used a multi-echo gradient-echo sequence (12 echoes; first echo time and echo spacing, 5 msec; repetition time, 2229 msec; flip angle, 30°; voxel size, 2 × 2 × 3 mm³; gap, 0.3 mm; 35 sections) with correction for macroscopic magnetic background field gradients by a sinc-Gauss excitation pulse (8,9) and half- and quarter-resolution k-space center acquisitions for motion correction (10). CBF was measured with pseudocontinuous arterial spin labeling (pCASL; label duration and post-labeling delay, 1800 msec; four background suppression pulses; two-dimensional echo-planar readout; echo time, 11 msec; repetition time, 4500 msec; 20 sections; voxel size, 3.28 × 3.5 × 6.0 $mm^3$; gap, 0.6 mm; 39 label-control pairs and a proton density-weighted M0 image), following the recommended implementation (11). Older-adults I and II used pCASL with a segmented three-dimensional GraSE readout instead (label duration, 1800 msec; post-labeling delay, 2000 msec; four background suppression pulses; echo time, 7.4 msec; repetition time, 4377 msec; flip angle, 90°; 16 sections; turbo spin-echo factor, 19; echo-planar imaging factor, 7; voxel size, 2.75 × 2.75 × 6.0 $mm^3$; three dynamics including a proton density-weighted M0 image; acquisition time, 5:41 min; 12). CBV was measured with dynamic susceptibility contrast MRI (single-shot gradient-echo echo-planar imaging; repetition time, 2000 msec; flip angle, 60°; voxel size, 2 × 2 × 3.5 $mm^3$; 35 sections; 80 dynamics), with a bolus of gadolinium-based contrast agent (0.1 mL/kg; 6–8 mL; 4 mL/s) after five dynamics, followed by 25 mL of saline (13). Anatomical images comprised a three-dimensional T1-weighted magnetization-prepared rapid gradient-echo image (repetition time, 9 msec; echo time, 4 msec; flip angle, 8°; 170 sections; voxel size, 1 × 1 × 1 $mm^3$) and a three-dimensional T2-weighted fluid-attenuated inversion recovery image to exclude brain lesions. In Young-adults I, BOLD fMRI was acquired with single-shot multiband echo-planar imaging (repetition time, 1200 msec; echo time, 30 msec; flip angle, 70°; voxel size, 3 × 3 × 3 $mm^3$; 40 sections; SENSE factor, 2; multiband factor, 2) together with a B0 field map. Rest and calculation conditions were presented in pseudorandomized order across the mqBOLD acquisitions (1). During the calculation task, participants extrapolated the step of a numerical sequence (n1, n2, n3, ?) and selected the missing value among three alternatives within 10 seconds; during rest, participants fixated a white cross on a black screen. The cross-vendor dataset was acquired on a 3-T Biograph mMR scanner (Siemens Healthineers, Erlangen, Germany) with a 12-channel head-neck coil; T2 mapping used a multi-echo two-dimensional turbo spin-echo sequence (8 echoes; first echo time and echo spacing, 16 msec; repetition time, 4870 msec), and the T2*, pCASL, and dynamic susceptibility contrast sequences were adapted to this platform (4).

**mqBOLD Processing and Analytical $CMRO_2$**

Quantitative parameter maps were calculated with in-house scripts in MATLAB and SPM12 (Wellcome Trust Centre for Human Neuroimaging, University College London, United Kingdom). T2 and T2* maps were obtained by monoexponential fits of the multi-echo spin- and gradient-echo data, corrected for macroscopic magnetic background fields and motion (6,7). The reversible transverse relaxation rate $R2' = 1/T2^* - 1/T2$ reflects the vascular deoxyhemoglobin content (14). Relative CBV was derived from the dynamic susceptibility contrast data by full integration of leakage-corrected $\Delta R2^*$ curves and normalized to a white matter value of 2.5% (13,15). OEF was calculated as $OEF = R2'/(c \cdot CBV)$ (5,16), with $c = \gamma \cdot 4/3 \cdot \pi \cdot \Delta\chi_0 \cdot hct \cdot B0$, where $\gamma = 2.675 \cdot 10^8\ s^{-1}\ T^{-1}$ is the gyromagnetic ratio, $\Delta\chi_0 = 0.264 \cdot 10^{-6}$ the susceptibility difference between fully deoxygenated and oxygenated blood, B0 = 3 T, and hct the small-vessel hematocrit, estimated as 85% of the individual large-vessel hematocrit (17); in Older-adults I, sex-specific reference hematocrit values were used (women, 0.40; men, 0.45; 18,19). CBF was calculated from the averaged pairwise differences of motion-corrected label and control images, scaled by the M0 image (11); on Philips scanners, CBF was increased by 25% to compensate for the signal attenuation by background suppression (12). Voxel-wise $CMRO_2$ was calculated with Fick's principle as $CMRO_2 = OEF \cdot CBF \cdot CaO_2$, where $CaO_2 = 0.334 \cdot hct \cdot 55.6 \cdot O_2sat$ is the arterial oxygen content and $O_2$sat the arterial oxygen saturation measured with pulse oximetry (20). All parameter maps were registered to the first echo of the T2 data. These analytical $CMRO_2$ maps, calculated in native space from all four mqBOLD inputs, served as the reference standard for training and evaluation.

**Preprocessing for Model Development**

All parameter maps were transformed into MNI152NLin2009cAsym space in two steps. First, maps were rigidly coregistered (6 degrees of freedom) to the native T1-weighted image with FSL FLIRT (version 6.0) after brain extraction with FSL BET, avoiding scaling or shearing that could bias quantitative values. Second, the participant-specific nonlinear transform from the T1-weighted image to MNI space, estimated with fMRIPrep (version 20.2.3), was applied with ANTs (version 2.3.3) at 2-mm isotropic resolution. To reduce memory and computation time, maps were resampled to 4-mm isotropic resolution and cropped to a common matrix of 48 × 56 × 48 voxels. Inputs and targets were restricted to the conservative cortical gray matter mask of Epp et al (1), because mqBOLD-derived OEF and $CMRO_2$ are biased in white matter and in areas with strong magnetic susceptibility effects, for example near air-tissue interfaces (7). We validated this pipeline against $CMRO_2$ maps calculated from parameter maps that were first resampled to 4 mm. Quantitative inputs (CBF, CBV, T2, and T2*) and the $CMRO_2$ target were kept in physical units. As nonquantitative inputs, we used the T1-weighted image and the degree centrality of resting-state fMRI, which reflects the number of functional connections of each voxel (21). Functional images were section-timing corrected, realigned, skull stripped, intensity normalized, cleaned from nuisance signals (scanner drift, head motion, and CompCor physiological noise), and band-pass filtered (0.01–0.1 Hz). Degree centrality was computed with AFNI 3dDegreeCentrality as the weighted sum of significant Pearson correlations ($P < .001$) between each voxel time series and all other gray matter voxels, smoothed (full width at half maximum, 6 mm), and transformed to the analysis grid. T1-weighted and degree centrality maps were z-scored within the gray matter mask with the statistics of the training participants (Table S1).

**Model Architecture and Training**

$CMRO_2$ was synthesized with a conditional generative adversarial network (GAN) for image-to-image translation (Fig. 1b; Fig. S1; Table S1; 22,23). The generator was a three-dimensional attention-residual U-Net (24–26) that received a variable number of input channels and returned a single-channel $CMRO_2$ map. The encoder comprised three stages of pre-activation residual blocks (batch normalization, LeakyReLU, and 3 × 3 × 3 convolution, with a 1 × 1 × 1 convolution shortcut) with 64, 128, and 256 feature channels, each followed by dropout (0.3, 0.3, and 0.4) and max pooling, and a bottleneck residual block with 512 channels and dropout (0.5). The decoder upsampled the features with transposed convolutions, weighted the encoder skip connections with attention gates before concatenation, and ended with a 1 × 1 × 1 convolution. The discriminator was a three-dimensional convolutional classifier conditioned on the generator inputs by channel-wise concatenation with either the analytical or the synthetic $CMRO_2$ map; it comprised three convolutional blocks (64, 128, and 256 channels; 3 × 3 × 3 kernels; LeakyReLU with slope 0.2; max pooling), global average pooling, and two fully connected layers returning a single logit. By judging input-output pairs, the discriminator favored synthetic maps that were consistent with the conditioning inputs.

The generator loss combined a voxel-wise L1 loss, a nonsaturating adversarial loss, and a structural similarity loss, $L = L1 + 0.01 \cdot Ladv + 0.5 \cdot (1 - SSIM)$. The discriminator was trained with binary cross-entropy to classify analytical pairs as real and synthetic pairs as synthetic. Both networks were optimized with Adam (learning rates, $1 \cdot 10^{-4}$ for the generator and $5 \cdot 10^{-5}$ for the discriminator) for 750 epochs with a batch size of 4, and the generator learning rate was reduced by a factor of 0.9 when the validation generator loss did not improve for 10 epochs. The architecture started from the U-Net of Liebert et al (27), and learning rates, dropout rates, and batch size were set in a preliminary study on training participants only; all input configurations used identical hyperparameters, none tuned on test data. Training used baseline rest data only, with fourfold participant-level cross-validation of the training set (random seed, 1234). Per fold, the checkpoint with the lowest validation generator loss was retained, and per model, the fold with the lowest validation generator loss was applied to all test data, without ensembling. Training data were augmented with random flips and 90° rotations applied identically to inputs and targets, and with Gaussian noise, gamma contrast, intensity shift, and bias field perturbations applied to inputs only, so that targets retained their physical values (Table S1).

We trained the full model with all four mqBOLD inputs and models with all seven combinations of CBF, T2, and T2* (three single inputs, three pairs, and one triplet). CBV was removed first because it requires contrast agent administration. For each number of inputs, we report the combination with the lowest cross-validated MAE (Table S3): the three-input model without CBV, the two-input CBF&T2* model, and the single-input CBF model. Additional single-input models used the T1-weighted image or degree centrality (16 training and 9 test participants) and were cross-validated in the same way. Because it matched the three-input model with one input fewer, the CBF&T2* model was used as the minimal-input model for all subsequent analyses.

### Model Evaluation

All evaluations were performed on test data within the gray matter mask and in physical units (μmol/100 g/min). We quantified the agreement between synthetic and analytical $CMRO_2$ with the voxel-wise Pearson correlation coefficient (r) and slope of the linear regression of synthetic on analytical values, and with the MAE, the mean absolute difference between synthetic and analytical $CMRO_2$. Group-level metrics were computed on the group median maps, that is, the voxel-wise median across participants (7961 gray matter voxels), with bias-corrected and accelerated bootstrap 95% CIs for r (2000 resamples); individual metrics were computed across the voxels of each participant. Voxels with non-positive analytical or synthetic $CMRO_2$ were excluded. Six Glasser parcels next to the petrous bone and the frontal sinus (TE1m, TE2a, TE1p, TE2p, a10p, and 11l, in both hemispheres; 194 of 8155 voxels) were excluded from all evaluations, because their analytical $CMRO_2$ in the test set was 2.5 to 12 median absolute deviations above the median cortical parcel, consistent with susceptibility-related overestimation of R2′; models were trained on the full mask. To relate the errors to the measurement noise of the analytical method, the within-session and between-day intraclass correlation coefficients of analytical $CMRO_2$ (0.89 and 0.61; 2) were converted into expected test-retest errors, $2/\sqrt{\pi} \cdot SD \cdot \sqrt{(1 - ICC)}$, where SD is the between-participant SD per voxel of each dataset (Table S2). Repeatability was assessed as the ICC(2,1) (two-way random effects, absolute agreement, single measurement; 28) of the participant-wise medians of the two within-session scans across 305 Glasser parcels with at least 5 mask voxels (29). Parcel-wise medians were used because correlated neighboring voxels would otherwise inflate the precision of voxel-wise estimates. The ICCs of each model were compared with those of analytical $CMRO_2$ with Wilcoxon signed-rank tests paired by parcel.

To test whether models trained on baseline rest data preserved task-evoked $CMRO_2$ changes, we applied them to the calculation task data of the Young-adults I test participants and identified the spatial patterns differentiating calculation from rest with mean-centered partial least squares analysis (30), separately for analytical and synthetic $CMRO_2$, using the pyls library. The significance of the latent variables was assessed with 3000 permutations and voxel reliability with 3000 bootstrap resamples; the sign of the latent variable was set so that positive bootstrap ratios indicate calculation > rest. Voxels with an absolute bootstrap ratio greater than 2, approximately a 95% CI, were considered reliable (31), and the overlap between the reliable analytical and synthetic patterns was quantified with the Dice coefficient, $2|X \cap Y|/(|X| + |Y|)$.

The minimal-input model was applied without retraining to the same-protocol and cross-vendor datasets. For the cross-vendor dataset, T2 maps derived from turbo spin-echo data replaced the GraSE-derived T2 maps used during training. Individual metrics of each dataset were compared with those of the test set with Games-Howell tests.

**Age Analyses**

Age effects were assessed in the test set ($n = 41$; 21–84 years) as the linear change per decade of median gray matter $CMRO_2$, with participant bootstrap 95% CIs (2000 resamples), and as the Pearson correlation with age. Differences between the synthetic and analytical declines were tested by regressing the participant-wise difference (synthetic minus analytical $CMRO_2$) on age, which is equivalent to the age-by-method interaction of a linear mixed model with a random intercept per participant. The participant-wise agreement metrics and the synthetic-minus-analytical difference were correlated with age (Table S4).

Because the model error depended on age, an uncorrected synthetic decline mixes the physiological age effect with this error, as known from brain-age models (32,33). We therefore report the uncorrected decline together with two corrections that correspond to two scenarios. Without analytical $CMRO_2$, as in new participants, the error can only be estimated in advance. For this age calibration, each of the 54 training participants, a validation participant in exactly one fold, was predicted by the model of that fold, so that their errors resemble those of new participants. In these predictions, the error (synthetic minus analytical median $CMRO_2$) was fitted as $a + b \cdot age$ ($a = 6.8$ μmol/100 g/min; $b = -0.08$ μmol/100 g/min per year; $P = .60$), and the calibrated synthetic $CMRO_2$ of each test participant was computed as synthetic $CMRO_2 - (a + b \cdot age)$, using only age. With analytical $CMRO_2$, as in method evaluation, the participant-wise regression slope between synthetic and analytical $CMRO_2$ is known and was included as a covariate ($CMRO_2 \sim age + slope$); the age coefficients of synthetic and analytical $CMRO_2$ and of their difference were compared as above, with participant bootstrap 95% CIs and P values from the regression t-test.

### Statistical Analysis

Individual metrics and parcel-wise ICCs were compared between models and between calculation and rest with Wilcoxon signed-rank tests, paired by participant or parcel, with Benjamini-Hochberg false discovery rate correction within each metric; the test set and the same-protocol and cross-vendor datasets were compared with Games-Howell tests. All tests were two-sided, and $P < .05$ indicated statistical significance. Analyses used Python (version 3.13.5), SciPy (version 1.16.1), scikit-learn (version 1.7.1), pyls (commit d8a19d5), and Nilearn (version 0.11.1); deep learning libraries, hardware, and training time are listed in Table S1. The code that reproduces all reported statistics and figures is available in the code repository (see Code Availability).

### Supplementary Figure 1

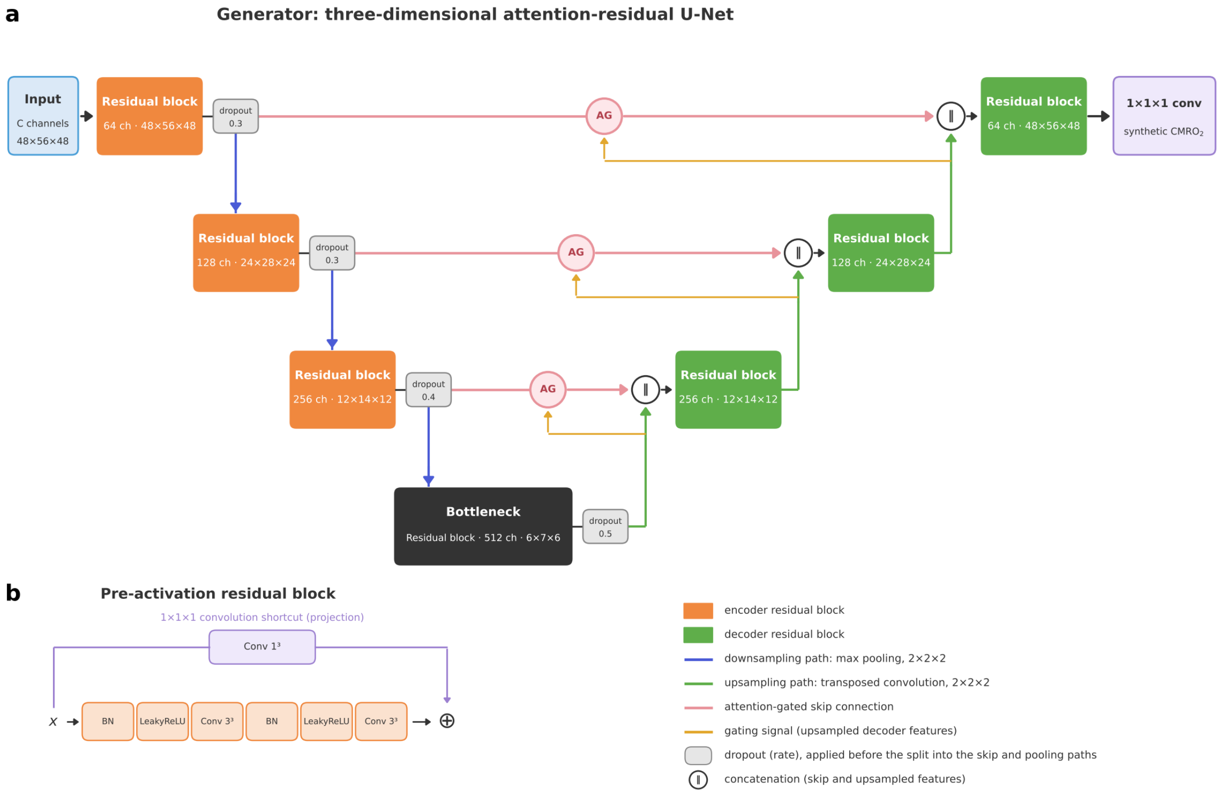

**Figure S1: Architecture of the generator**. (a) The generator is a three-dimensional U-Net with three encoder stages, a bottleneck, and three decoder stages; numbers indicate feature channels and spatial dimensions (voxels, 4-mm isotropic). Dropout follows each encoder residual block and the bottleneck, before the split into the pooling path and the skip connection. Attention gates (26) weight the encoder features by voxel-wise coefficients computed from the upsampled decoder features before concatenation. (b) Each stage uses a pre-activation residual block with a 1 × 1 × 1 convolution shortcut, because every block changes the number of channels. The discriminator is specified in Table S1. AG = attention gate, BN = batch normalization, C = number of input maps, $CMRO_2$ = cerebral metabolic rate of oxygen.

### Supplementary Figure 2

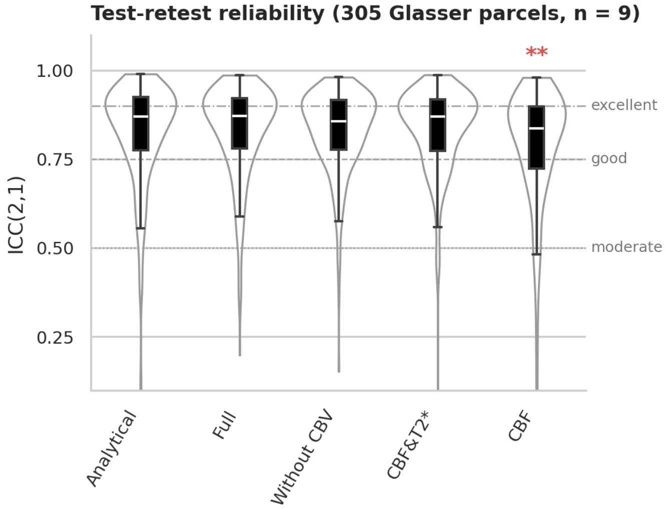

**Figure S2: Test-retest reliability of analytical and synthetic $CMRO_2$.** Intraclass correlation coefficients (ICC(2,1); two-way random effects, absolute agreement, single measurement) between two resting-state scans acquired within one session in nine Young-adults II test participants, computed per Glasser parcel from participant-wise parcel medians (305 parcels with at least five gray matter mask voxels). Violins show the distribution across parcels, boxes the interquartile range, and white lines the median. Horizontal lines mark the thresholds for moderate (0.5), good (0.75), and excellent (0.9) reliability (34). ** $P < .01$, model versus analytical $CMRO_2$ (Wilcoxon signed-rank test paired by parcel, false discovery rate corrected across models); three parcels of the CBF model and one of the CBF&T2* model with ICC < 0.1 lie below the axis.

### Supplementary Table 1
#### Table S1: Network Architecture and Training Hyperparameters

| Component | Stage or parameter | Specification | Output size |
|---|---|---|---|
| Generator | Encoder 1 | Residual block, C → 64 channels; dropout, 0.3; max pooling, 2 × 2 × 2 (stride 2) | 64 × 48 × 56 × 48 |
| Generator | Encoder 2 | Residual block, 64 → 128 channels; dropout, 0.3; max pooling, 2 × 2 × 2 (stride 2) | 128 × 24 × 28 × 24 |
| Generator | Encoder 3 | Residual block, 128 → 256 channels; dropout, 0.4; max pooling, 2 × 2 × 2 (stride 2) | 256 × 12 × 14 × 12 |
| Generator | Bottleneck | Residual block, 256 → 512 channels; dropout, 0.5 | 512 × 6 × 7 × 6 |
| Generator | Decoder 1 | Transposed convolution, 512 → 256 (2 × 2 × 2, stride 2); attention gate (gating, upsampled features; skip, encoder 3); concatenation, 512 channels; residual block, 512 → 256 | 256 × 12 × 14 × 12 |
| Generator | Decoder 2 | Transposed convolution, 256 → 128; attention gate (skip, encoder 2); concatenation, 256 channels; residual block, 256 → 128 | 128 × 24 × 28 × 24 |
| Generator | Decoder 3 | Transposed convolution, 128 → 64; attention gate (skip, encoder 1); concatenation, 128 channels; residual block, 128 → 64 | 64 × 48 × 56 × 48 |
| Generator | Output | 1 × 1 × 1 convolution, 64 → 1 channel (synthetic $CMRO_2$) | 1 × 48 × 56 × 48 |
| Generator | Residual block | Pre-activation: two units of batch normalization, LeakyReLU (slope 0.01), and 3 × 3 × 3 convolution, added to a 1 × 1 × 1 convolution shortcut (projection; every block changes the channel number) | |
| Discriminator | Input | Concatenation of the C input maps and the analytical or synthetic $CMRO_2$ map | (C + 1) × 48 × 56 × 48 |
| Discriminator | Block 1 | 3 × 3 × 3 convolution (padding 1), C + 1 → 64; LeakyReLU (slope 0.2); max pooling, 2 × 2 × 2 | 64 × 24 × 28 × 24 |
| Discriminator | Block 2 | 3 × 3 × 3 convolution, 64 → 128; LeakyReLU (0.2); max pooling | 128 × 12 × 14 × 12 |
| Discriminator | Block 3 | 3 × 3 × 3 convolution, 128 → 256; LeakyReLU (0.2); max pooling | 256 × 6 × 7 × 6 |
| Discriminator | Classifier head | Global average pooling; fully connected, 256 → 1024, LeakyReLU (0.2); fully connected, 1024 → 1 | 1 logit |
| Losses | Generator loss | L1 + 0.01 · adversarial (nonsaturating binary cross-entropy with logits) + 0.5 · (1 − SSIM); SSIM data range from the analytical map | |
| Losses | Discriminator loss | $0.5 \cdot BCE(D[x, y], 1) + 0.5 \cdot BCE(D[x, \hat{y}], 0)$ | |
| Optimization | Optimizer | Adam; learning rate, $1 \cdot 10^{-4}$ (generator) and $5 \cdot 10^{-5}$ (discriminator) | |
| Optimization | Learning rate schedule | Generator learning rate × 0.9 after 10 epochs without improvement of the validation generator loss | |

| | | |
|---|---|---|
| Optimization | Epochs; batch size | 750; 4 |
| Optimization | Hyperparameter selection | Starting point: the U-Net of Liebert et al (27), with three encoder and three decoder stages around a bottleneck, 64 to 512 feature channels, batch normalization and LeakyReLU, dropout in the deeper stages, an L1 plus SSIM loss, and the checkpoint with the lowest validation loss; extended here to three dimensions with residual blocks, attention gates, and an adversarial loss. Learning rates, dropout rates, and batch size were then set empirically for stable adversarial training in a preliminary study on training participants only (unpublished data). As in Liebert et al (27), one independent network was trained per input configuration with identical hyperparameters, none tuned on test data |
| Model selection | Cross-validation and checkpoints | Fourfold participant-level cross-validation within the training set (random seed 1234); per fold, checkpoint with lowest validation generator loss retained; fold with the lowest validation generator loss applied to all test data, no ensembling |
| Augmentation | Inputs and targets | Random flips along each axis (probability, 0.5 per axis); random 90° rotations (probability, 0.5; up to three quarter turns) |
| Augmentation | Inputs only | Applied after input scaling, that is, to quantitative maps in physical units: Gaussian noise, SD = 0.05; gamma contrast adjustment, $\gamma$ = 0.9–1.1; intensity shift, offset ≤ 0.1; bias field (probability, 0.5 each) |
| Input scaling | All inputs | Quantitative maps in physical units; T1-weighted image z-scored over nonzero gray matter voxels before augmentation, with statistics of the fold's training participants during cross-validation and of all 54 training participants at test time; degree centrality z-scored in the same way (Young-adults I fMRI subset: fold training participants or all 16 training participants); all maps multiplied by the gray matter mask |
| Reproducibility | Initialization and seeds | PyTorch defaults; fixed random seed (1234) for Python, NumPy, and PyTorch, including CUDA |
| Software and hardware | Software | Python 3.13.5; PyTorch 2.7.0 (CUDA 11.8); MONAI 1.4.0; pytorch-msssim 1.0.0 |
| Software and hardware | Hardware | NVIDIA GeForce RTX 4090 GPU (24 GB) |
| Software and hardware | Training time | About 12 s per epoch and 2.5 h per fold (10 h per four-fold model), with two models trained in parallel on one GPU |

Note: Output sizes are given as channels × voxels (4-mm isotropic). With two inputs, the generator has 22.8 million trainable parameters and the discriminator 1.4 million. C = number of input maps (one to four quantitative maps, or the T1-weighted image or degree centrality map). BCE = binary cross-entropy, $CMRO_2$ = cerebral metabolic rate of oxygen, D = discriminator, SSIM = structural similarity index measure, x = input maps, y = analytical $CMRO_2$ map, $\hat{y}$ = synthetic $CMRO_2$ map. Dropout follows each encoder residual block and thus acts on both the pooling path and the skip connection to the decoder.

### Supplementary Table 2

#### Table S2: Model Error Relative to the Test-Retest Error of Analytical $CMRO_2$

| Figure | Dataset (n) | Model | MAE, median [IQR] | Retest error, within / between session | MAE ÷ retest error, within / between | Parcel level: MAE ÷ retest error, within / between |
|---|---|---|---|---|---|---|
| 2 | Test set, rest (41) | Full | 25.7 [20.4–30.6] | 24.7 / 46.5 | 1.04 / 0.55 | 1.05 / 0.56 |
| 2 | Test set, rest (41) | Without CBV | 38.1 [30.2–43.8] | 24.7 / 46.5 | 1.54 / 0.82 | 1.46 / 0.77 |
| 2 | Test set, rest (41) | CBF&T2* | 38.1 [30.8–46.0] | 24.7 / 46.5 | 1.54 / 0.82 | 1.53 / 0.81 |
| 2 | Test set, rest (41) | CBF | 48.8 [41.0–58.9] | 24.7 / 46.5 | 1.97 / 1.05 | 1.92 / 1.02 |
| 2 | Test set, rest (41) | T1-weighted | 67.9 [58.6–81.4] | 24.7 / 46.5 | 2.75 / 1.46 | 2.74 / 1.45 |
| 2 | Test set (fMRI subset), rest (9) | fMRI | 78.2 [69.8–90.7] | 18.9 / 35.6 | 4.14 / 2.20 | 3.39 / 1.80 |
| 3 | Calculation task (10) | Full | 25.3 [23.2–28.0] | 19.1 / 35.9 | 1.32 / 0.70 | 1.32 / 0.70 |
| 3 | Calculation task (10) | Without CBV | 36.5 [33.5–42.4] | 19.1 / 35.9 | 1.91 / 1.02 | 1.74 / 0.92 |
| 3 | Calculation task (10) | CBF&T2* | 38.0 [35.4–44.8] | 19.1 / 35.9 | 1.99 / 1.06 | 1.81 / 0.96 |
| 3 | Calculation task (10) | CBF | 48.6 [44.3–56.1] | 19.1 / 35.9 | 2.54 / 1.35 | 2.36 / 1.26 |
| 4 | Same-protocol (24) | CBF&T2* | 43.0 [38.8–50.2] | 22.3 / 41.9 | 1.93 / 1.02 | 1.97 / 1.04 |
| 4 | Cross-vendor (12) | CBF&T2* | 58.9 [52.3–70.8] | 28.0 / 52.8 | 2.10 / 1.12 | 1.80 / 0.96 |

Note: MAE is the median [interquartile range] of the participant-wise mean absolute errors between synthetic and analytical $CMRO_2$ across gray matter voxels (μmol/100 g/min), as shown in Figures 2–4. The retest error is the expected mean absolute difference between two analytical $CMRO_2$ measurements, $2/\sqrt{\pi} \cdot SD \cdot \sqrt{(1 - ICC)}$, where SD is the between-participant SD of analytical $CMRO_2$ per voxel in each dataset (median across voxels) and ICC is the within-session (0.89) or between-day (0.61) intraclass correlation coefficient of analytical $CMRO_2$ across Glasser parcels (2). The voxel-level benchmark assumes that the parcel-level ICC also holds at the voxel level. Parcel level: the same ratios with MAE and SD computed on Glasser parcel medians (parcels with at least five voxels), the level at which the ICC was estimated. Ratios below 1 indicate a model error smaller than the test-retest error. ICC = intraclass correlation coefficient, MAE = mean absolute error.

### Supplementary Table 3

#### Table S3: Cross-validated Performance and Selection of Input Configurations

| Inputs | No. of inputs | Validation generator loss | MAE (µmol/100 g/min) | r | Slope | Selected fold | Selected |
|---|---|---|---|---|---|---|---|
| **CBV, CBF, T2, T2* (full model)** | **4** | **2.21 ± 0.20** | **30.9 ± 4.3** | **0.91 ± 0.02** | **0.78 ± 0.06** | **1** | **Yes** |
| **CBF, T2, T2* (without CBV)** | **3** | **3.08 ± 0.22** | **43.2 ± 3.1** | **0.80 ± 0.03** | **0.63 ± 0.04** | **3** | **Yes** |
| **CBF, T2* (CBF&T2*)** | **2** | **3.22 ± 0.15** | **44.1 ± 2.4** | **0.78 ± 0.02** | **0.60 ± 0.03** | **1** | **Yes** |
| CBF, T2 | 2 | 4.23 ± 0.23 | 57.1 ± 2.9 | 0.60 ± 0.06 | 0.31 ± 0.08 | 3 | |
| T2, T2* | 2 | 4.11 ± 0.50 | 59.2 ± 6.4 | 0.66 ± 0.05 | 0.44 ± 0.06 | 1 | |
| **CBF** | **1** | **4.22 ± 0.28** | **58.0 ± 3.4** | **0.61 ± 0.09** | **0.31 ± 0.08** | **3** | **Yes** |
| T2* | 1 | 4.73 ± 0.38 | 67.3 ± 5.2 | 0.41 ± 0.10 | 0.23 ± 0.09 | 2 | |
| T2 | 1 | 5.35 ± 0.25 | 75.6 ± 2.3 | 0.24 ± 0.23 | 0.09 ± 0.09 | 3 | |
| T1-weighted | 1 | 6.09 ± 0.49 | 79.0 ± 9.2 | 0.17 ± 0.08 | 0.03 ± 0.02 | 2 | |
| Degree centrality (resting-state fMRI) | 1 | 6.66 ± 0.42 | 92.5 ± 10.1 | 0.31 ± 0.06 | 0.18 ± 0.07 | 3 | |

Note: Data are means ± SDs across the four cross-validation folds of the main cohort training set (n = 54; 12–14 validation participants per fold); test participants were not used. For each fold, the saved generator (the one with the lowest validation loss during training) was applied to the fold's validation participants without augmentation. Validation generator loss = L1 + 0.01 · adversarial + 0.5 · (1 − SSIM), as during training. MAE, r, and slope were computed per participant across gray matter voxels (excluding non-positive values) and summarized as the median per fold. For each number of inputs, the configuration with the lowest cross-validated MAE was selected (in bold); for each configuration, the fold with the lowest validation generator loss was applied to all test datasets. The T1-weighted and degree centrality models were predefined comparisons and were not part of the input reduction; the degree centrality model was cross-validated within the 16 Young-adults I training participants with resting-state fMRI (four validation participants per fold). MAE = mean absolute error, SSIM = structural similarity index measure.

### Supplementary Table 4

**Table S4: Association of Participant-wise Agreement with Age in the Test Set**

| Model | r | Slope | MAE | Relative MAE | Median bias | Median synthetic $CMRO_2$ |
|---|---|---|---|---|---|---|
| CBV, CBF, T2, T2* (full model) | −0.58 (< .001) | −0.78 (< .001) | 0.11 (.48) | 0.41 (.007) | −0.68 (< .001) | −0.63 (< .001) |
| CBF, T2, T2* (without CBV) | −0.35 (.03) | −0.32 (.04) | −0.19 (.25) | 0.33 (.04) | −0.15 (.35) | −0.48 (.002) |
| **CBF, T2* (minimal input)** | **−0.51 (< .001)** | **−0.31 (.047)** | **−0.31 (.048)** | **0.10 (.52)** | **−0.37 (.02)** | **−0.55 (< .001)** |
| CBF | −0.28 (.08) | −0.01 (.97) | −0.20 (.21) | 0.20 (.22) | −0.39 (.01) | −0.71 (< .001) |
| T1-weighted | 0.17 (.29) | −0.14 (.37) | −0.51 (< .001) | −0.32 (.04) | 0.42 (.006) | 0.30 (.055) |

Note: Data are Pearson correlation coefficients with age and, in parentheses, uncorrected P values (n = 41 test participants, 21–84 years). Agreement measures were computed per participant across gray matter voxels as in Figure 2h–2j: r and slope of synthetic versus analytical $CMRO_2$ and MAE (μmol/100 g/min). Relative MAE = MAE divided by the participant's median analytical $CMRO_2$; median bias = median synthetic minus median analytical $CMRO_2$. For comparison, median analytical $CMRO_2$ correlated with age (r = −0.42; P = .007). A slope, bias, or relative MAE that changes with age could inflate or mask the age association of synthetic $CMRO_2$; for the minimal-input CBF&T2* model, slope and median bias decreased with age, consistent with its steeper age decline (see Results). MAE = mean absolute error.
